# Association of polyunsaturated fatty acids with cholelithiasis risk and the role of plasma lipid mediators: insights from NHANES 2017-2020 and Mendelian randomization

**DOI:** 10.1101/2024.07.22.24310765

**Authors:** Yuxuan Chen, Wei Chen, Jin Qian, Xuanchen Jin, Congying Wang, Yihu Zheng

## Abstract

**Background & aims:** Previous studies have suggested a potential link between polyunsaturated fatty acid (PUFA) intake and the risk of cholelithiasis. Omega-3 fatty acids, a key subfamily of PUFAs, have been identified in observational studies as playing a role in lipid regulation and potentially serving as a protective factor against cholelithiasis. In this study, we aim to investigate this association further by analyzing data from the 2017-2020 National Health and Nutrition Examination Survey (NHANES) and conducting Mendelian randomization (MR) analyses.

**Methods:** We employed weighted multivariate-adjusted logistic regression analyses to examine the association between PUFAs and cholelithiasis risk using data from NHANES 2017-2020. Additionally, a two-sample Mendelian randomization (MR) study was conducted utilizing pooled data from Genome-Wide Association Studies (GWAS) to establish the causal relationship between PUFAs and cholelithiasis. Following this, we performed two-step MR mediation analyses to investigate the mediating role of plasma lipids in the pathway, focusing on the strongly positive subfamily of PUFAs, Omega-3, in relation to plasma circulating lipids and cholelithiasis.

**Results:** Our observational study in NHANES included 7,527 participants. Weighted multivariate-adjusted logistic regression analyses initially revealed a negative association between PUFAs, their subclasses, and cholelithiasis. However, this association became nonsignificant after adjusting for multiple covariates. In contrast, MR analyses identified a significant negative association between PUFAs (OR=0.75 [95% CI, 0.58∼0.98]) and Omega-3 (OR=0.79 [95% CI, 0.7∼0.9]) and the risk of cholelithiasis. Specifically, Omega-3 was associated with a reduced risk of developing cholelithiasis (OR=0.77 [95% CI, 0.65∼0.91]), possibly due to the upregulation of LDL-C levels (Beta=0.24 [95% CI, 0.1∼0.38]). This upregulation of LDL-C subsequently lowered the risk of cholelithiasis (OR=0.77 [95% CI, 0.65∼0.91]), with the mediating effect of LDL-C accounting for 28% of the overall association.

**Conclusions:** Both cross-sectional observational analyses and Mendelian randomization (MR) analyses demonstrated a negative correlation between polyunsaturated fatty acids (PUFAs) and cholelithiasis. Omega-3 fatty acids seem to play a key role in this association by increasing plasma LDL-C levels, which in turn may help reduce the risk of cholelithiasis.

## 1. Introduction

Cholelithiasis, the most common disease of the biliary system, affects people worldwide. Its development is influenced by genetic and environmental factors, and the exact causes and mechanisms remain incompletely understood. Gaining a better understanding of these factors is crucial for the prevention and treatment of cholelithiasis.

Unsaturated fatty acids, essential to the human body, are divided into monounsaturated fatty acids (MUFAs) and polyunsaturated fatty acids (PUFAs) based on the number and location of double bonds. Among PUFAs, Omega-3 (ω-3) and Omega-6 (ω-6) are the primary families relevant to human health. Over the past decades, numerous studies have highlighted their impact on hypertension, cardiovascular disease, type 2 diabetes, arthritis, Alzheimer’s disease, depression, infection resistance, and inflammation regulation [1, 2]. There is also growing evidence that PUFAs might influence hepatobiliary diseases through specific metabolic pathways.

Animal experiments have shown that polyunsaturated fats can inhibit cholesterol stone formation [3, 4]. Epidemiological studies on the relationship between PUFAs and gallstone disease have produced mixed results. For instance, a prospective cohort study found that high consumption of polyunsaturated fats reduced the risk of gallstone disease in men [5]. Conversely, another study found no direct association between PUFA intake and gallstone disease [6].

Omega-3 fatty acids, a significant branch of PUFAs, cannot be synthesized naturally by the human body and must be obtained through diet [7]. These fatty acids play an important role in biliary tract health. Clinical studies have shown that dietary supplementation with Omega-3 can reduce bile cholesterol saturation in patients with gallstones, and additionally, combining Omega-3 with ursodeoxycholic acid (UDCA), a first-line treatment alternative to gallbladder surgery, has improved therapeutic outcomes [8–10]. Early animal studies also demonstrated that Omega-3 fatty acids could reduce cholesterol excretion and lower plasma cholesterol levels by increasing cholesterol transport to the bile [11, 12].

Among the various risk factors for cholelithiasis, plasma lipid levels are particularly influential. Previous studies have highlighted a significant link between gallstones and abnormal plasma lipid levels [13]. This raises important questions about the interplay between PUFAs, plasma lipids, and cholelithiasis risk. Specifically, it is crucial to determine whether plasma lipids mediate the relationship between PUFAs and cholelithiasis and to understand their role in this process. This area of research holds significant implications for both clinical disease management and mechanistic studies of gallstone formation. However, detailed investigations into these associations are still lacking.

Existing observational studies have limitations, such as small sample sizes, insufficient adjustment for key variables, and incomplete assessment of different PUFA subtypes. The National Health and Nutrition Examination Survey (NHANES) offers a valuable resource in this context. As an ongoing, comprehensive cross-sectional study conducted by the National Center for Health Statistics (NCHS), NHANES provides large, high-quality, and representative data on the U.S. population, making it ideal for examining the relationship between PUFAs and gallstone disease.

While cross-sectional studies can be confounded by various factors that are challenging to control, Mendelian randomization (MR) offers a robust method for assessing causal relationships using genetic information. MR leverages genetic variants, such as single nucleotide polymorphisms (SNPs), to determine causal links between exposures and outcomes, minimizing the impact of confounding variables [14]. MR analyses can also be used to investigate mediators, further reducing potential biases from uncontrolled confounders [15]. Given the inconsistencies in previous observational studies and the need for more definitive causal evidence, MR provides a valuable complement to observational research.

This research aims to investigate the potential association between PUFAs and cholelithiasis, focusing on the possible mediating role of plasma lipids. We will use both cross-sectional analyses and MR methods, utilizing data from the NHANES database to enhance our understanding of these relationships.

## 2. Material and methods

### 2.1. NHANES Transect Study

#### 2.1.1. NHANES Study Design and Participants

Figure 1 provides an overview of our cross-sectional observational study. The data for this analysis were sourced from the publicly accessible NHANES database (https://www.cdc.gov/nchs/nhanes/index.htm). The NHANES study protocol was approved by the NCHS Ethics Review Board, and informed consent was obtained from all participants. Since our study utilized de-identified, publicly available data, no additional institutional review board approval was required.

**Figure 1.**
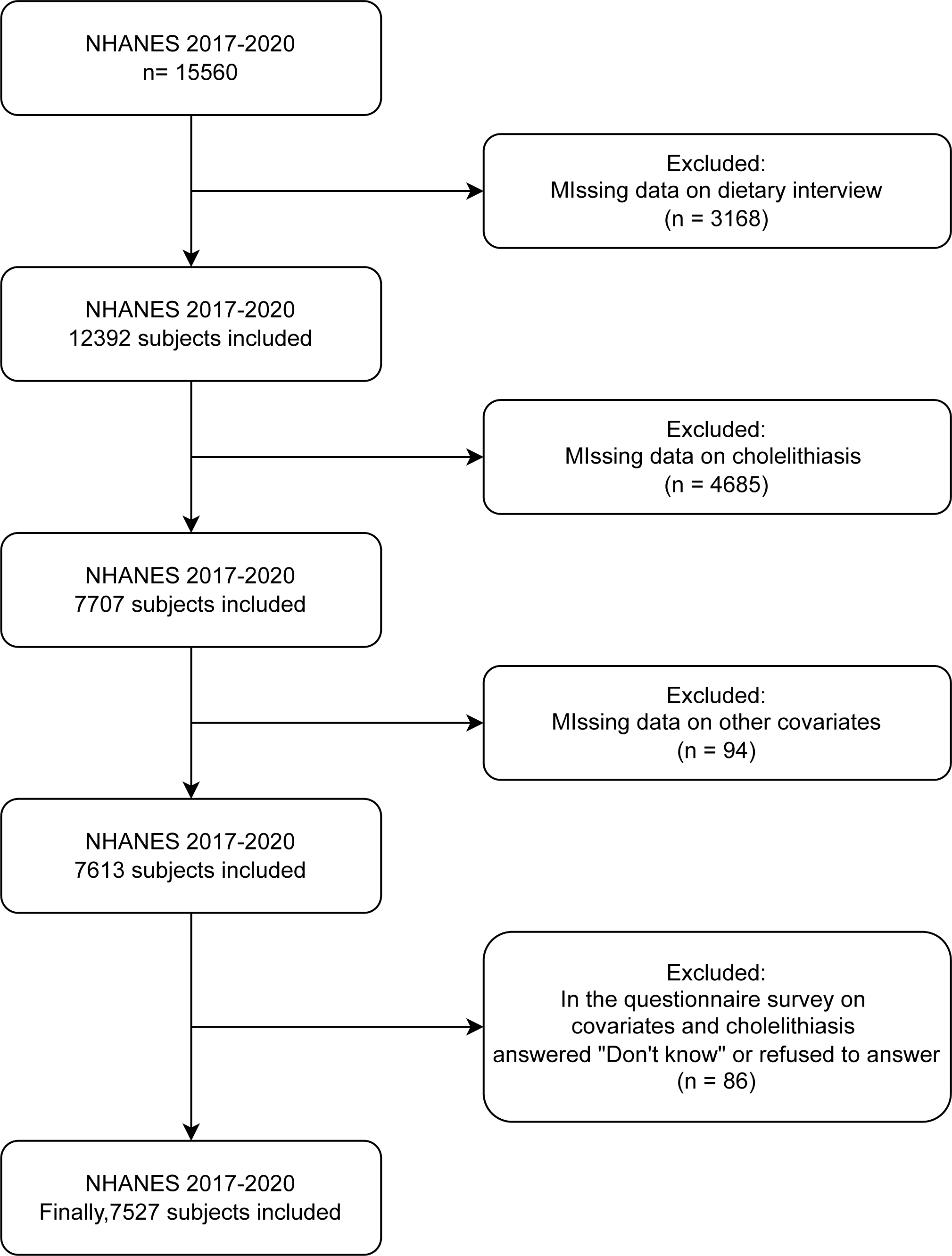
NHANES cross sectional study design.

We analyzed data from before the 2017-2020 pandemic, as cholelithiasis-related information was only available during this period. The NHANES 2017-2020 dataset initially included 15,560 participants. Our study focused on individuals who completed the Dietary 24-Hour Recall Survey during this time. We excluded: (1) 3,168 individuals who did not complete the Dietary 24-Hour Recall Survey and Gallstone Questionnaire, (2) 4,685 individuals with missing data from the Gallstone Questionnaire, (3) 94 individuals with incomplete data on relevant covariates, and (4) 86 individuals who provided unsatisfactory responses (e.g., “don’t know” or “refused to answer”) to the dietary recall and related covariate questionnaires. This resulted in a final study sample of 7,527 participants.

#### 2.1.2. Dietary Unsaturated Fatty Acid Intake and Definition of Cholelithiasis as Used in NHANES

The outcome of this study was the presence or absence of gallstone disease, determined using the 2017-2020 NHANES Health Questionnaire. Participants were asked, “Has a doctor or other health professional ever told (YOU/SP)(YOU/SHE/HIM) that you have gallstones?“

The major exposure was the total daily intake of dietary PUFAs, measured in grams. Participants underwent two dietary 24-hour recalls on two separate interview days, reporting the type and amount of food and beverages consumed in the 24 hours prior to the interview (midnight to midnight). This data was used to estimate energy (calories) intake and the intake of nutrients and other components.

The dietary interview component, known as What We Eat in America (WWEIA), is a collaboration between the U.S. Department of Agriculture (USDA) and the U.S. Department of Health and Human Services (DHHS). USDA Food and Nutritional Database for Dietary Studies (FNDDS) codes were used to identify foods and beverages in the survey. FNDDS 2017-2018 and FNDDS 2019-2020 were used to address intake reported in the 2017-2020 sample. The mean total intake of PUFAs for each participant over the two interview days was used in this study.

#### 2.1.3. Other covariates used in NHANES

In this analysis, we considered age, race, gender, education, body mass index (BMI), smoking status, hypertension, hyperlipidemia, and diabetes mellitus as potential confounders. Educational attainment was classified into two groups: high school and below, and above high school. Race was categorized as non-Hispanic white, non-Hispanic black, Mexican American, non-Hispanic Asian, other Hispanic, and other race/multiracial. BMI was calculated based on height and weight measurements collected by technicians at the Mobile Examination Center (MEC). Smoking status was derived from a questionnaire, with participants considered smokers if they had smoked at least 100 cigarettes in their lifetime. Hypertension and hyperlipidemia were classified as either present or absent, while diabetes mellitus was categorized as present, absent, or borderline. These conditions were determined based on questionnaire responses, with participants considered positive if they had ever been diagnosed with any of these conditions by a health professional.

#### 2.1.4. Statistical analyses in NHANES

In our cross-sectional study, we employed multivariable-adjusted logistic regression to evaluate the association between PUFAs and gallstone disease. We assessed three covariate-adjusted models: Model 1: Unadjusted for covariates. Model 2: Adjusted for age, race, education level, and BMI. Model 3: Adjusted for all covariates. Given that NHANES uses a complex multistage sampling design, where the sample is divided into four levels (counties, segments, households, and individuals) with varying probabilities of selection at each stage, the data are not independent. To account for this multi-stage probability sampling design, we weighted and adjusted the data to produce estimates representative of the U.S. population. All analyses were based on weighted estimates using sample weights provided by NHANES.

Statistical analyses were performed using R version 4.3.1 (http://www.r-project.org), with the “survey” and “gtsummary” packages. Continuous variables were described using means and 95% confidence intervals (CIs), while categorical variables were described using survey percentages. A P value of less than 0.05 was considered statistically significant.

### 2.2. Mendelian randomization studies

#### 2.2.1. MR Study Design

Figure 2 provides an overview of our current Mendelian randomization (MR) research. To obtain valid causal estimates, MR models must satisfy three basic assumptions: 1) Association Assumption: There is a strong correlation between the instrumental variable and the exposure factor. 2) Exclusivity Assumption: The instrumental variable is uncorrelated with potential confounders. 3) Independence Assumption: The instrumental variable affects the outcome only through the exposure. Our study was conducted in two steps. First, we used a two-sample Mendelian randomization (TSMR) method to identify the exposure variable as PUFAs and the outcome variable as cholelithiasis. We further screened for strong positive exposure to Omega-3 using a two-step Mendelian randomization model to verify whether plasma lipids were a potential mediator in the causal relationship between Omega-3 and cholelithiasis and to calculate the associated mediating effects.

**Figure 2.**
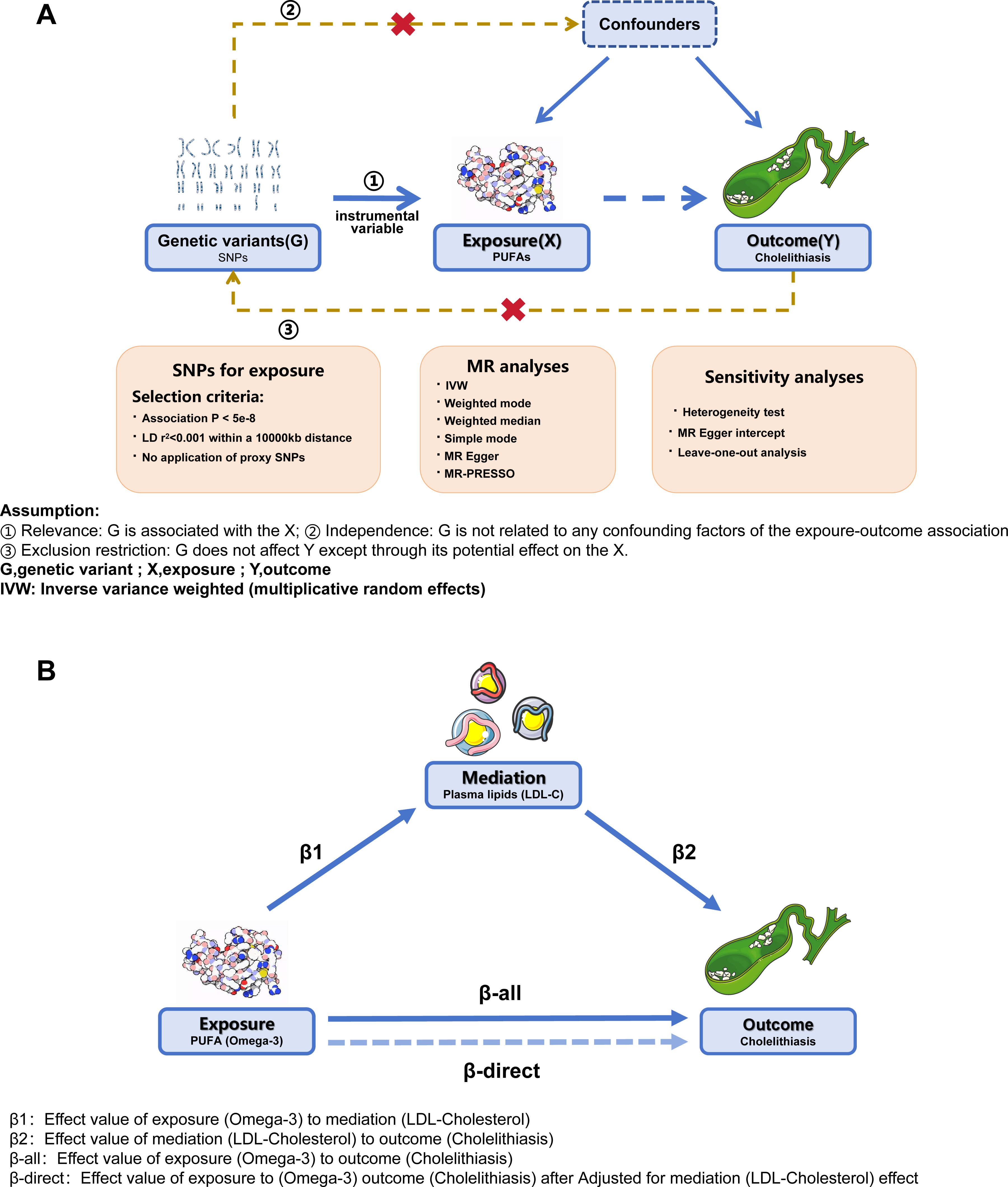
Assumptions of the Two-sample Mendelian randomization (A) and Two-step mediated Mendelian randomization (B) study design.

#### 2.2.2. Data sources for MR analysis

Our MR analyses utilized publicly available statistics from large-scale Genome-Wide Association Studies (GWAS) datasets. Summary-level data on genetic variants associated with PUFAs exposure were obtained from the UK Biobank (https://www.ukbiobank.ac.uk/), which included 114,999 European ethnic samples. GWAS summary data for genetic variants associated with cholelithiasis as an outcome endpoint were obtained from the FinnGen Consortium’s Round 10 study (https://www.finngen.fi/en), which included 40,191 samples. For genetic variation data on plasma lipids as a mediating variable, we used the largest available multi-GWAS meta-data from the Global Lipids Genetics Consortium (https://csg.sph.umich.edu/willer/public/glgc-lipids2021/), which included a total of 132,016 samples of European ethnicity. To avoid potential sample overlap issues with the PUFAs data, we used a meta dataset with the UK Biobank cohort removed for this part of the study (Table 1) [16].

**Table 1:** Characteristic of GWAS employed in the MR study.

| **Items** | **GWAS ID** | **Consortium** | **Sample size** | **Population** |
| --- | --- | --- | --- | --- |
| PUFA | met-d-PUFA | UK Biobank | 114,999 | European |
| MUFA | met-d-MUFA | UK Biobank | 114,999 | European |
| Omega-3 | met-d-Omega_3 | UK Biobank | 114,999 | European |
| Omega-6 | met-d-Omega_6 | UK Biobank | 114,999 | European |
| HDL-C (without UKB cohort) | / | GLGC | 1,320,016 | European |
| LDL-C (without UKB cohort) | / | GLGC | 1,320,016 | European |
| Triglyceride (without UKB cohort) | / | GLGC | 1,320,016 | European |
| Cholelithiasis | K11_CHOLELITH | FinnGen | 401,832 | European |

#### 2.2.3. Selection of Genetic Instrumental Variables

To identify genetic variants for estimating the causal relationship between exposure (PUFAs, plasma lipids) and cholelithiasis, we set stringent filtering conditions. We applied a genome-wide significance level of P < 5*10^-8^, to screen for genetic variants strongly associated with exposure. Additionally, we excluded single nucleotide polymorphisms (SNPs) with linkage disequilibrium properties (r^2^ < 0.001, 10,000 kb). To avoid potential bias from weak instrumental variables, we selected genetic variants with F-statistics > 10 for follow-up analyses.

#### 2.2.4. Two-sample MR

We used an inverse variance weighted (IVW) random effects model [17] to evaluate the results of genetic prediction. Additionally, four complementary methods, MR Egger, Simple mode, Weighted mode, and Weighted median [18, 19], were employed as supplementary approaches to the IVW method. We provided the odds ratios (or beta) and 95% confidence intervals for all five methods. The IVW method was the primary tool for assessing causal associations between exposures and outcomes. Bidirectional Mendelian randomization was used to determine the directionality of causality. Furthermore, the MR-PRESSO method [20] was applied as a complementary approach to the primary IVW outcome, enhancing the robustness of positive results when statistically significant (P < 0.05).

#### 2.2.5. MR mediation analysis

Based on the two-sample Mendelian randomization (TSMR), exposures that were validated to be statistically significant (P < 0.05) under the IVW method were further analyzed with plasma lipids as the outcome for MR analysis. This approach allowed us to construct a two-step MR mediation analysis model. In this model, the plasma lipid outcome with a positive screening result was used as an exposure in further TSMR analysis with cholelithiasis as the outcome. Subsequently, the mediator ratio was calculated.

The above method was used to obtain the β1 value for PUFAs to plasma lipids, the β2 value in the MR analysis of plasma lipid exposure to cholelithiasis outcome after excluding PUFA-related SNPs, and the β-all value for the total effect of exposure to PUFAs to cholelithiasis outcome, with a focus on whether the mechanism could be passed. “β3=β1*β2” indicates the indirect effect of the mediator in the mechanism. “β-direct” indicates the direct effect of the mediator in the mechanism.

#### 2.2.6. Sensitivity analysis

Because inverse variance weighted (IVW) estimates may be biased by introducing pleiotropic instrumental variables, we conducted sensitivity analyses to detect pleiotropy in causal estimates. We used MR-Egger’s intercept to estimate the horizontal pleiotropy of genetic variation, considering P < 0.05 as an indication of potential horizontal pleiotropy. Cochrane’s Q test was employed to assess potential heterogeneity. If the P value was greater than 0.05, indicating no evidence of heterogeneity, we used the fixed-effects IVW method as the main analysis. Conversely, if significant heterogeneity (P < 0.05) was present, we applied the random-effects IVW method. Additionally, we performed leave-one-out analyses to determine whether the results were influenced by individual variants. Funnel plots were used to check for the presence of heterogeneity among individual genetic variants. In the absence of heterogeneity, the funnel plots should exhibit a symmetrical shape.

#### 2.2.7. Statistical analyses in MR

All MR analyses were performed using R version 4.3.1 (http://www.r-project.org). The ‘TwosampleMR’ and ‘MR-PRESSO’ packages were used for the analyses, while the ‘ggplot’ package was utilized for statistical plotting. P values <0.05 were considered statistically significant.

## 3. Results

### 3.1. Population characteristics of study subjects according to cholelithiasis in NHANES

Table 2 displays the demographic, clinical, and dietary characteristics of the study participants, who were divided into 804 cholelithiasis patients and 6,723 non-cholelithiasis patients based on the presence of cholelithiasis. Cholelithiasis patients were older, predominantly female, and non-Hispanic white. There were no significant differences in educational attainment between the two groups. However, the cholelithiasis patients had a higher BMI and a greater tendency to smoke. Additionally, the cholelithiasis group exhibited higher rates of hypertension, hyperlipidemia, and diabetes compared to the non-cholelithiasis group. Regarding dietary characteristics, the cholelithiasis group consumed less unsaturated fatty acids, monounsaturated fatty acids, polyunsaturated fatty acids, Octadecadienoic acid (PUFA 18:2), Eicosatetraenoic acid (PUFA 20:4), Eicosapentaenoic acid (PUFA 20:5), and Docosapentaenoic acid (PUFA 22:5). However, there was no significant difference between the two groups in terms of levels of Octadecatrienoic acid (PUFA 18:3), Octadecatetraenoic acid (PUFA 18:4), and Docosahexaenoic acid (PUFA 22:6).

**Table 2:** Baseline characteristics of participants.

| Characteristic | N<sup>1</sup> | Overall, N = 7527 (100%)<sup>2</sup> | Cholelithiasis |  | P Value<sup>3</sup> |
| --- | --- | --- | --- | --- | --- |
|  |  |  | Yes, N = 804 (9.9%)<sup>2</sup> | No, N = 6723 (90%)<sup>2</sup> |  |
| **Sex** | 7,527 |  |  |  | **<0.001** |
| *female* |  | 3,876 (46%) | 573 (67%) | 3,303 (44%) |  |
| *male* |  | 3,651 (54%) | 231 (33%) | 3,420 (56%) |  |
| **Age (years)** | 7,527 | 50.0 (34.0, 63.0) | 59.0 (45.0, 70.0) | 49.0 (33.0, 62.0) | **<0.001** |
| **Race** | 7,527 |  |  |  | **0.001** |
| *Non-Hispanic White* |  | 2,691 (36%) | 345 (43%) | 2,346 (36%) |  |
| *Non-Hispanic Black* |  | 2,016 (29%) | 166 (23%) | 1,850 (29%) |  |
| *Mexican American* |  | 870 (12%) | 96 (11%) | 774 (12%) |  |
| *Non-Hispanic Asian* |  | 834 (9.4%) | 50 (6.3%) | 784 (9.8%) |  |
| *Other Hispanic* |  | 759 (9.2%) | 94 (11%) | 665 (9.0%) |  |
| *Other Race / Multi-Racial* |  | 357 (4.9%) | 53 (6.2%) | 304 (4.7%) |  |
| **Education level** | 7,527 |  |  |  | **>0.9** |
| *High school or less* |  | 3,122 (39%) | 342 (39%) | 2,780 (39%) |  |
| *More than high school* |  | 4,405 (61%) | 462 (61%) | 3,943 (61%) |  |
| **BMI** | 7,527 | 29 (25, 34) | 32 (28, 38) | 29 (25, 34) | **<0.001** |
| **Smoke** | 7,527 | 3,180 (43%) | 375 (48%) | 2,805 (43%) | **0.02** |
| **Hypertension** | 7,527 | 2,873 (37%) | 443 (55%) | 2,430 (35%) | **<0.001** |
| **Hyperlipidemia** | 7,527 | 2,701 (35%) | 400 (52%) | 2,301 (33%) | **<0.001** |
| **Diabetes** | 7,527 |  |  |  | **<0.001** |
| *Yes* |  | 1,129 (15%) | 206 (26%) | 923 (13%) |  |
| *No* |  | 6,190 (83%) | 576 (70%) | 5,614 (84%) |  |
| *Borderline* |  | 208 (2.9%) | 22 (4.1%) | 186 (2.8%) |  |
| **Saturated fatty acid<sup>4</sup>** | 7,527 | 29 (21, 40) | 28 (20, 38) | 29 (21, 40) | **0.016** |
| **Monounsaturated fatty acid<sup>4</sup>** | 7,527 | 32 (23, 44) | 30 (22, 40) | 33 (24, 44) | **<0.001** |
| **Polyunsaturated fatty acid<sup>4</sup>** | 7,527 | 23 (17, 32) | 22 (16, 32) | 24 (17, 32) | **0.024** |
| **Octadecadienoic acid (PUFA 18:2)<sup>4</sup>** | 7,527 | 21 (15, 29) | 19 (14, 28) | 21 (15, 29) | **0.022** |
| **Octadecatrenoic acid (PUFA 18:3)<sup>4</sup>** | 7,527 | 2.10 (1.43, 2.98) | 2.06 (1.41, 2.94) | 2.11 (1.43, 2.99) | 0.2 |
| **Octadecatetraenoic acid (PUFA 18:4)<sup>4</sup>** | 7,527 | 0.002 (0.001, 0.005) | 0.002 (0.001, 0.005) | 0.002 (0.001, 0.006) | 0.13 |
| **Eicosatetraenoic acid (PUFA 20:4)<sup>4</sup>** | 7,527 | 0.15 (0.09, 0.22) | 0.13 (0.08, 0.20) | 0.15 (0.09, 0.23) | **<0.001** |
| **Eicosapentaenoic acid (PUFA 20:5)<sup>4</sup>** | 7,527 | 0.01 (0.01, 0.02) | 0.01 (0.01, 0.02) | 0.01 (0.01, 0.02) | **0.032** |
| **Docosapentaenoic acid (PUFA 22:5)<sup>4</sup>** | 7,527 | 0.022 (0.013, 0.036) | 0.020 (0.012, 0.030) | 0.023 (0.013, 0.036) | **<0.001** |
| **Docosahexaenoic acid (PUFA 22:6)<sup>4</sup>** | 7,527 | 0.02 (0.01, 0.07) | 0.02 (0.00, 0.06) | 0.02 (0.01, 0.07) | 0.4 |

<sup>1</sup>N not Missing (unweighted)<sup>2</sup>median (IQR) for continuous; n (%) for categorical<sup>3</sup>chi-squared test with Rao & Scott's second-order correction; Wilcoxon rank-sum test for complex survey samples<sup>4</sup>The unit of measurement is gram (gm)

### 3.2. Observational associations between PUFAs and cholelithiasis in NHANES

The results of multifactorial regression analyses demonstrated a correlation between polyunsaturated fatty acids (PUFAs) and the risk of cholelithiasis (Table 3). In Model 1, unsaturated fatty acids (OR=0.99 [95% CI, 0.99∼1.00]), monounsaturated fatty acids (OR=0.99 [95% CI, 0.98– 1.00]), polyunsaturated fatty acids (OR=0.99 [95% CI, 0.98∼1.00]), PUFA 18:2 (OR=0.99 [95% CI, 0.98∼1.00]), PUFA 18:3 (OR=0.93 [95% CI, 0.88∼0.99]), and PUFA 20:4 (OR=0.20 [95% CI, 0.10∼0.43]) were significantly and negatively associated with cholelithiasis risk. After adjusting for relevant covariates in Model 2, monounsaturated fatty acids (OR=0.99 [95% CI, 0.98∼1.00]), polyunsaturated fatty acids (OR=0.99 [95% CI, 0.98∼1.00]), PUFA 18:2 (OR=0.99 [95% CI, 0.98∼1.00]), PUFA 18:3 (OR=0.93 [95% CI, 0.88∼0.98]), and PUFA 20:4 (OR=0.21 [95% CI, 0.10∼0.43]) remained significantly and negatively associated with cholelithiasis risk. However, after further adjustment for additional covariates in Model 3, these significant associations became non-significant. Only PUFA 20:4 (OR=0.53 [95% CI, 0.25∼1.12]) continued to show a negative association with the development of cholelithiasis, although this was not statistically significant (p=0.088).

**Table 3:** Association between PUFAs subgroups and cholelithiasis.

| **Group** | **Characteristic** | **OR<sup>1</sup>** | **95% CI<sup>1</sup>** | **p-value** |
| --- | --- | --- | --- | --- |
| **modle1** |  |  |  |  |
|  | Saturated fatty acid | 0.99 | 0.99, 1.00 | **0.021** |
|  | Monounsaturated fatty acid | 0.99 | 0.98, 1.00 | **0.007** |
|  | Polyunsaturated fatty acid | 0.99 | 0.98, 1.00 | **0.015** |
|  | Octadecadienoic acid (PUFA 18:2) | 0.99 | 0.98, 1.00 | **0.017** |
|  | Octadecadienoic acid (PUFA 18:3) | 0.93 | 0.88, 0.99 | **0.019** |
|  | Octadecatetraenoic acid (PUFA 18:4) | 0.11 | 0.00, 4.13 | 0.2 |
|  | Eicosatetraenoic acid (PUFA 20:4) | 0.2 | 0.10, 0.43 | **<0.001** |
|  | Eicosapentaenoic acid (PUFA 20:5) | 0.64 | 0.31, 1.32 | 0.2 |
|  | Docosapentaenoic acid (PUFA 22:5) | 0.05 | 0.00, 3.42 | 0.2 |
|  | Docosahexaenoic acid (PUFA 22:6) | 0.66 | 0.43, 1.01 | 0.057 |
| **modle2** |  |  |  |  |
|  | Saturated fatty acid | 0.99 | 0.99, 1.00 | 0.081 |
|  | Monounsaturated fatty acid | 0.99 | 0.98, 1.00 | **0.022** |
|  | Polyunsaturated fatty acid | 0.99 | 0.98, 1.00 | **0.036** |
|  | Octadecadienoic acid (PUFA 18:2) | 0.99 | 0.98, 1.00 | **0.048** |
|  | Octadecadienoic acid (PUFA 18:3) | 0.93 | 0.88, 0.98 | **0.013** |
|  | Eicosatetraenoic acid (PUFA 20:4) | 0.21 | 0.10, 0.43 | **<0.001** |
| **modle3** |  |  |  |  |
|  | Monounsaturated fatty acid | 1 | 0.99, 1.01 | >0.9 |
|  | Polyunsaturated fatty acid | 1 | 0.99, 1.01 | 0.8 |
|  | Octadecadienoic acid (PUFA 18:2) | 1 | 0.99, 1.01 | 0.9 |
|  | Octadecadienoic acid (PUFA 18:3) | 0.97 | 0.92, 1.02 | 0.2 |
|  | Eicosatetraenoic acid (PUFA 20:4) | 0.53 | 0.25, 1.12 | 0.088 |

<sup>1</sup>OR = Odds Ratio, CI = Confidence Interval
Modle1: Unadjusted for covariates
Modle2: Adjusted for age, race, education level and BMI
Modle3: Adjusted for all covariates

### 3.3. Causal relationships between PUFAs and cholelithiasis risk in two-sample MR

Since the multivariate regression analysis found some negative correlation between PUFAs and cholelithiasis risk, we concluded that PUFAs might be a protective factor against cholelithiasis. Therefore, we further performed Mendelian randomization (MR) analysis to infer the causal relationship between PUFAs and cholelithiasis risk. As shown in Table 4, using the inverse variance weighted (IVW) method, the results indicated that PUFAs (OR=0.75 [95% CI, 0.58∼0.98]) were negatively associated with cholelithiasis risk. Among them, Omega-3 (OR=0.79 [95% CI, 0.7∼0.9]) was significantly associated with a reduced risk of developing cholelithiasis, with a strong positive result (p=0.00037). In addition, the results of the other complementary methods were consistent in the direction of causal estimation (all OR<1), and the MR-PRESSO test also yielded a positive result. This evidence suggests that the findings are reliably robust.

**Table 4:** MR estimates from each method with cholelithiasis as the outcome.

| Exposure | MR method | nSNPs | Beta | SE | OR (95% CI) | P value |
| --- | --- | --- | --- | --- | --- | --- |
| PUFA | IVW | 53 | -0.28 | 0.13 | 0.75 (0.58,0.98) | 0.032 |
|  | MR Egger | 53 | -0.43 | 0.27 | 0.65 (0.39,1.1) | 0.116 |
|  | Simple mode | 53 | -0.01 | 0.07 | 0.99 (0.86,1.14) | 0.930 |
|  | Weighted mode | 53 | -0.12 | 0.05 | 0.89 (0.81,0.99) | 0.029 |
|  | Weighted median | 53 | -0.15 | 0.04 | 0.86 (0.79,0.94) | 0.001 |
|  | MR-PRESSO | 53 | -0.28 | 0.14 | 0.75 (0.58,0.98) | 0.037 |
| MUFA | IVW | 55 | 0.01 | 0.09 | 1.01 (0.84,1.21) | 0.933 |
|  | MR Egger | 55 | -0.25 | 0.17 | 0.78 (0.56,1.08) | 0.142 |
|  | Simple mode | 55 | 0.04 | 0.09 | 1.04 (0.88,1.23) | 0.655 |
|  | Weighted mode | 55 | -0.01 | 0.07 | 0.99 (0.87,1.13) | 0.879 |
|  | Weighted median | 55 | -0.04 | 0.05 | 0.96 (0.87,1.06) | 0.403 |
|  | MR-PRESSO | 55 | 0.01 | 0.09 | 1.01 (0.84,1.21) | 0.933 |
| Omega-3 | IVW | 45 | -0.23 | 0.06 | 0.79 (0.7,0.9) | 3.71E-04 |
|  | MR Egger | 45 | -0.19 | 0.12 | 0.83 (0.66,1.05) | 0.126 |
|  | Simple mode | 45 | -0.14 | 0.08 | 0.87 (0.74,1.02) | 0.085 |
|  | Weighted mode | 45 | -0.11 | 0.04 | 0.9 (0.82,0.98) | 0.018 |
|  | Weighted median | 45 | -0.14 | 0.05 | 0.87 (0.79,0.95) | 0.002 |
|  | MR-PRESSO | 45 | -0.23 | 0.07 | 0.79 (0.69,0.91) | 0.001 |
| Omega-6 | IVW | 53 | -0.23 | 0.14 | 0.79 (0.6,1.04) | 0.099 |
|  | MR Egger | 53 | -0.29 | 0.27 | 0.74 (0.44,1.27) | 0.283 |
|  | Simple mode | 53 | -0.07 | 0.07 | 0.93 (0.82,1.06) | 0.291 |
|  | Weighted mode | 53 | -0.15 | 0.05 | 0.86 (0.79,0.94) | 0.002 |
|  | Weighted median | 53 | -0.17 | 0.04 | 0.85 (0.78,0.92) | 7.51E-05 |
|  | MR-PRESSO | 53 | -0.23 | 0.14 | 0.79 (0.6,1.05) | 0.105 |

MR: Mendelian randomization; SE: Standard error; OR: Odds ratio; CI: Confidence interval
PRESSO: Pleiotropy residuals sum and outlier; IVW: Inverse variance weighted (multiplicative random effects)
PUFA: Polyunsaturated fatty acid; MUFA: Monounsaturated fatty acids

Figure 3 is a forest plot depicting the estimated effect of exposure on the risk of cholelithiasis as measured by different MR methods.

**Figure 3.**
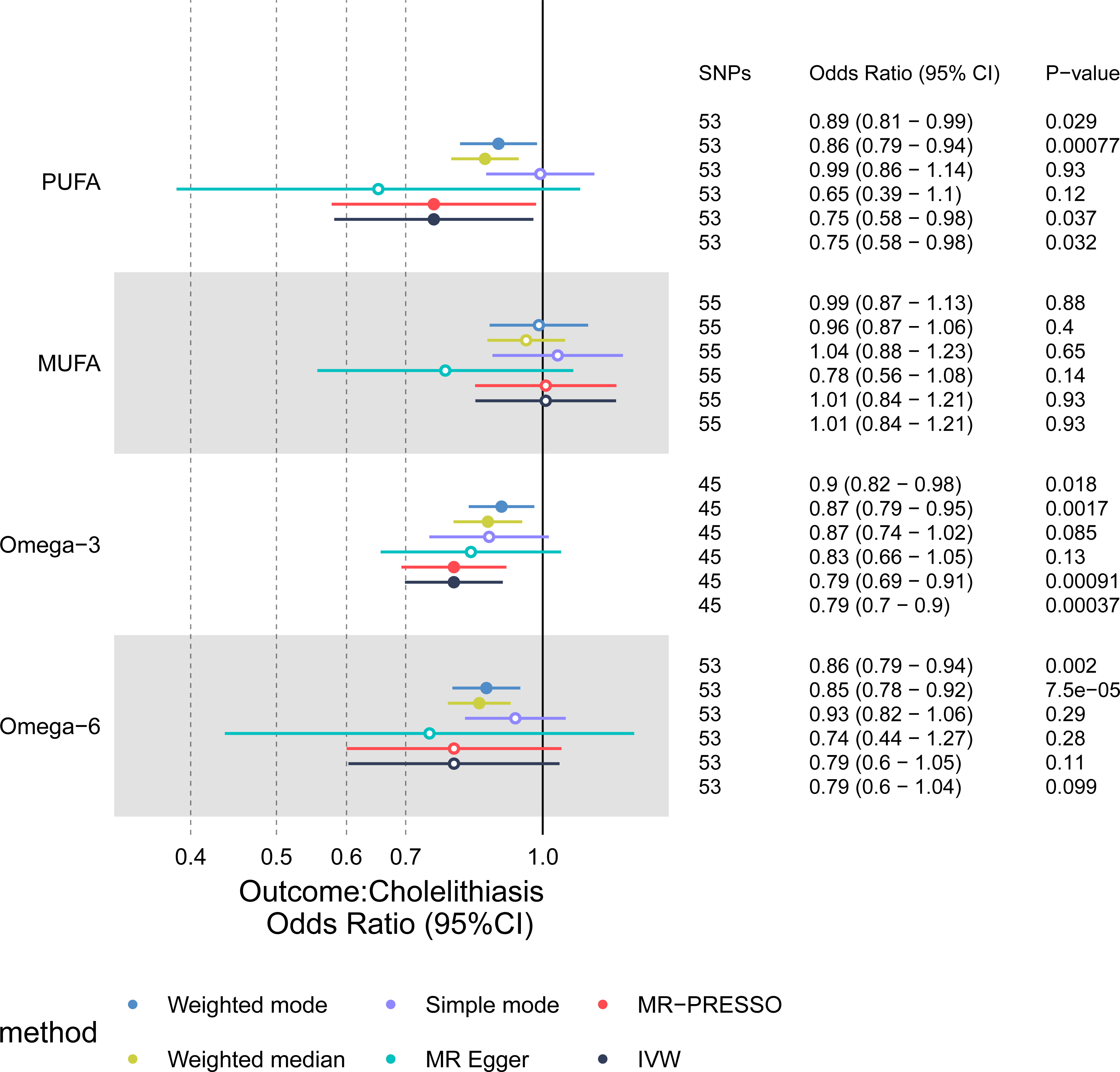
Forest plot shows the effects of PUFAs and MUFAs on cholelithiasis using different methods. CI: Confidence interval. Solid dots indicate significance with p < 0.05. Hollow circles indicate non-significance with p ≥ 0.05.

Notably, we observed potential SNP heterogeneity (Q-pval<0.05) in the effect of exposure on cholelithiasis risk (Supplementary Table 1), and thus we chose to apply a multiplicative random-effects IVW approach in estimating the causal effect, assessing the effect size between exposure and cholelithiasis risk by adjusting for the heterogeneity of the measurements. To assess whether exposure influenced the development of cholelithiasis through other potential pathways, we further performed a horizontal multivariate analysis. The results showed that no pleiotropy was detected for any of the four exposures after MR Egger’s intercept test (all P>0.05, Supplementary Table 2) The symmetry of the funnel plot indicated the same result (Supplementary Table 3), and the leave-one-out analysis further confirmed that the causality between PUFAs and cholelithiasis development was not driven by any single SNP (Supplementary Table 4). Thus, the above results indicate that our findings are stable and reliable.

### 3.4. LDL-C mediated Omega-3 as a protective factor in cholelithiasis

Based on these results, we further explored the deep intrinsic link between the strongly positive association (p=0.00037) of Omega-3 and the risk of cholelithiasis. Omega-3 was used as an exposure, and three plasma circulating lipids (HDL cholesterol (HDL-C), LDL cholesterol (LDL-C), and triglycerides) were used as exposures for MR analysis.

As shown in Table 5 (Figure 4), under the primary IVW MR analysis method, we observed a positive correlation between Omega-3 and LDL-C (Beta = 0.24 [95% CI, 0.1∼0.38]), and a positive correlation between Omega-3 and triglycerides (Beta = 0.29 [95% CI, 0.09∼0.49]). The other MR analysis methods were consistent in the direction of the causal effect between Omega-3 and LDL-C (all Beta > 0). Similarly, we did not observe potential horizontal pleiotropy in sensitivity analyses (all MR Egger endings P > 0.05, Supplementary Table 2), and funnel plots and leave-one-out analyses did not reveal significant abnormalities (Supplementary Table 3, 4).

**Figure 4.**
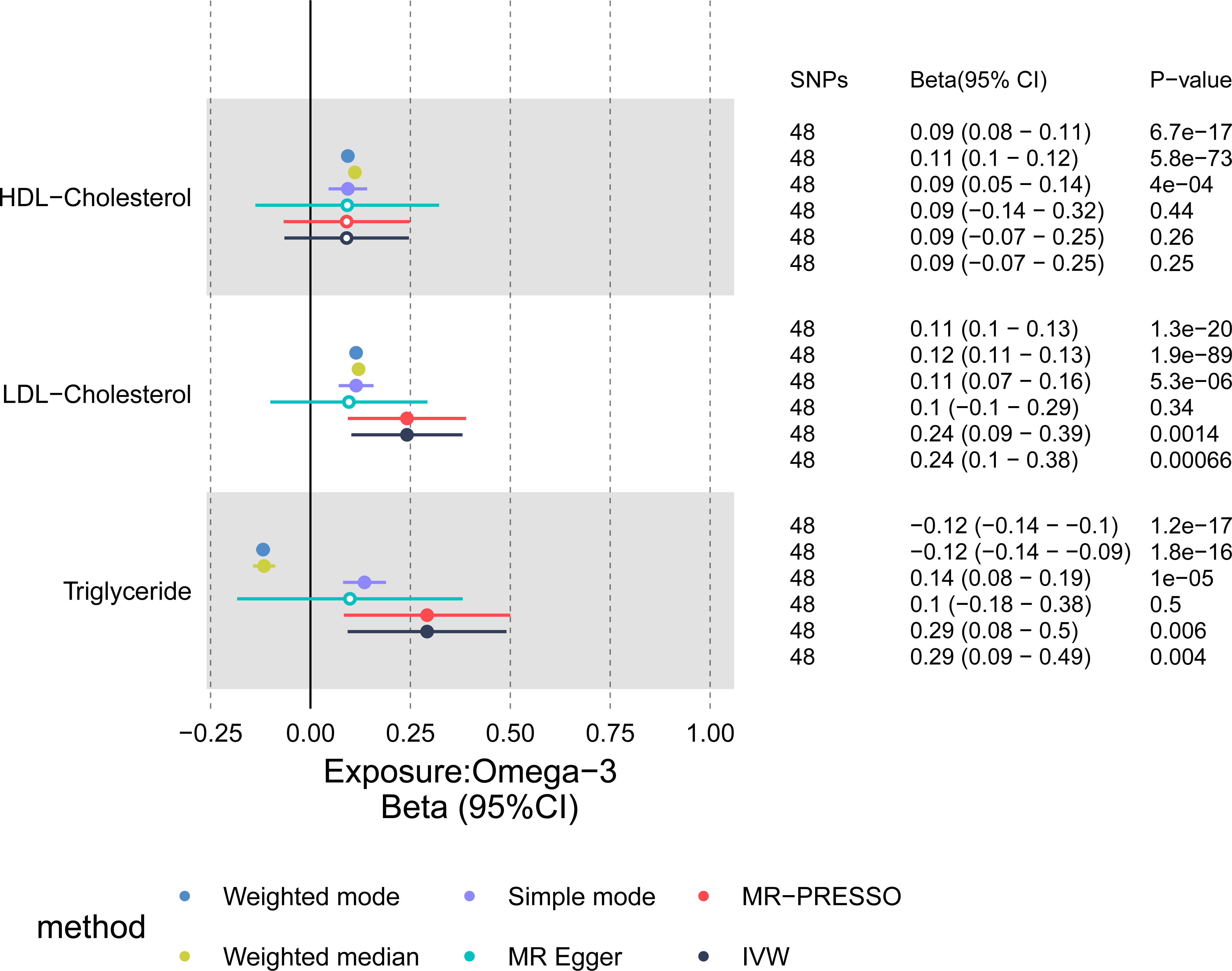
Forest plot shows the effects of plasma lipids on Omega-3 using different methods. CI: Confidence interval. Solid dots indicate significance with p < 0.05. Hollow circles indicate non-significance with p ≥ 0.05.

**Table 5:** MR estimates from each method with Omega-3 as the exposure.

| **Outcome** | **MR method** | **nSNPs** | **Beta (95% CI)** | **SE** | **OR** | **P value** |
| --- | --- | --- | --- | --- | --- | --- |
| HDL-Cholesterol | IVW | 48 | 0.09 (-0.07,0.25) | 0.08 | 1.09 | 0.255 |
|  | MR Egger | 48 | 0.09 (-0.14,0.32) | 0.12 | 1.10 | 0.437 |
|  | Simple mode | 48 | 0.09 (0.05,0.14) | 0.02 | 1.10 | 4.04E-04 |
|  | Weighted mode | 48 | 0.09 (0.08,0.11) | 0.01 | 1.10 | 6.70E-17 |
|  | Weighted median | 48 | 0.11 (0.1,0.12) | 0.01 | 1.12 | 5.78E-73 |
|  | MR-PRESSO | 48 | 0.09 (-0.07,0.25) | 0.08 | 1.09 | 0.261 |
| LDL-Cholesterol | IVW | 48 | 0.24 (0.1,0.38) | 0.07 | 1.27 | 0.001 |
|  | MR Egger | 48 | 0.1 (-0.1,0.29) | 0.10 | 1.10 | 0.342 |
|  | Simple mode | 48 | 0.11 (0.07,0.16) | 0.02 | 1.12 | 5.28E-06 |
|  | Weighted mode | 48 | 0.11 (0.1,0.13) | 0.01 | 1.12 | 1.31E-20 |
|  | Weighted median | 48 | 0.12 (0.11,0.13) | 0.01 | 1.13 | 1.91E-89 |
|  | MR-PRESSO | 48 | 0.24 (0.09,0.39) | 0.08 | 1.27 | 0.001 |
| Triglyceride | IVW | 48 | 0.29 (0.09,0.49) | 0.10 | 1.34 | 0.004 |
|  | MR Egger | 48 | 0.1 (-0.18,0.38) | 0.14 | 1.10 | 0.496 |
|  | Simple mode | 48 | 0.14 (0.08,0.19) | 0.03 | 1.14 | 1.05E-05 |
|  | Weighted mode | 48 | -0.12 (-0.14,-0.1) | 0.01 | 0.89 | 1.20E-17 |
|  | Weighted median | 48 | -0.12 (-0.14,-0.09) | 0.01 | 0.89 | 1.82E-16 |
|  | MR-PRESSO | 48 | 0.29 (0.08,0.5) | 0.11 | 1.34 | 0.006 |

MR: Mendelian randomization; SE: Standard error; OR: Odds ratio; CI: Confidence interval
PRESSO: Pleiotropy residuals sum and outlier; IVW: Inverse variance weighted (multiplicative random effects)

Further, we performed MR analyses of LDL-C and triglycerides as exposures and cholelithiasis as the outcome. As shown in Figure 5, LDL-C was negatively associated with the risk of cholelithiasis under the IVW method (OR=0.77 [95% CI, 0.65∼0.91]), and the same direction of effect was maintained for the other complementary MR analysis methods (all OR < 1). No horizontal pleiotropy was detected in the sensitivity analyses (Supplementary Table 2), and funnel plots and leave-one-out analyses supported our conclusions (Supplementary Table 3, 4).

**Figure 5.**
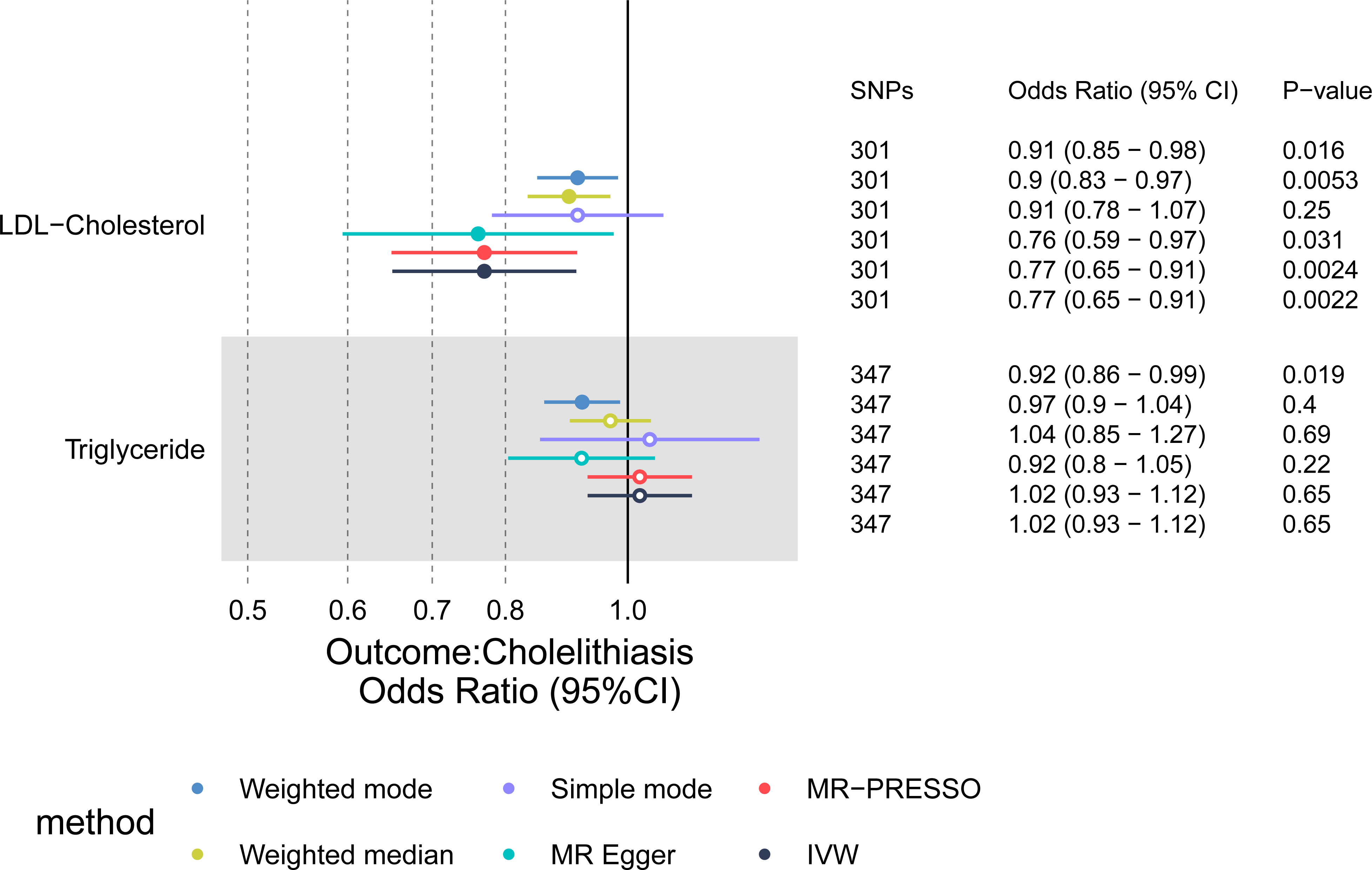
Forest plot shows the effects of plasma lipids on cholelithiasis using different methods. CI: Confidence interval. Solid dots indicate significance with p < 0.05. Hollow circles indicate non-significance with p ≥ 0.05.

To calculate the mediation effect size, 3 SNPs (rs9987289, rs34707604, rs58542926) obtained in the analysis of Omega-3 as exposure to LDL-C as the outcome were excluded. An MR analysis of LDL-C as exposure and cholelithiasis as the outcome after SNP exclusion showed a negative correlation (OR=0.76 [95% CI, 0.64∼0.91]). An effect value of β2 = −0.268 was obtained. Using the IVW method as the gold standard and Omega-3 as the exposure with LDL-C as the outcome, β1 = 0.241 was obtained. The total effect of Omega-3 and the risk of cholelithiasis, β-all = −0.231. The mediating effect, β3 = β1*β2 = −0.065, and the direct effect, β-direct = β-all - β3 = −0.167, 95% CI [−0.093∼(−0.036)]. The mediating effect share (β3/β-all) was calculated to be 28% (0.279) [95% CI, 0.154∼0.405]. These results are summarized in Table 6.

**Table 6:** MR of the association between plasma lipids and cholelithiasis.

| **Exposure** | **MR method** | **nSNPs** | **Beta** | **SE** | **OR (95% CI)** | **P value** |
| --- | --- | --- | --- | --- | --- | --- |
| LDL-Cholesterol | IVW | 301 | -0.26 | 0.09 | 0.77 (0.65,0.91) | 0.002 |
|  | MR Egger | 301 | -0.27 | 0.13 | 0.76 (0.59,0.97) | 0.031 |
|  | Simple mode | 301 | -0.09 | 0.08 | 0.91 (0.78,1.07) | 0.253 |
|  | Weighted mode | 301 | -0.09 | 0.04 | 0.91 (0.85,0.98) | 0.016 |
|  | Weighted median | 301 | -0.11 | 0.04 | 0.9 (0.83,0.97) | 0.005 |
|  | MR-PRESSO | 301 | -0.26 | 0.09 | 0.77 (0.65,0.91) | 0.002 |
| Triglyceride | IVW | 347 | 0.02 | 0.05 | 1.02 (0.93,1.12) | 0.651 |
|  | MR Egger | 347 | -0.08 | 0.07 | 0.92 (0.8,1.05) | 0.218 |
|  | Simple mode | 347 | 0.04 | 0.10 | 1.04 (0.85,1.27) | 0.695 |
|  | Weighted mode | 347 | -0.08 | 0.04 | 0.92 (0.86,0.99) | 0.019 |
|  | Weighted median | 347 | -0.03 | 0.04 | 0.97 (0.9,1.04) | 0.400 |
|  | MR-PRESSO | 347 | 0.02 | 0.05 | 1.02 (0.93,1.12) | 0.651 |

MR: Mendelian randomization; SE: Standard error; OR: Odds ratio; CI: Confidence interval
PRESSO: Pleiotropy residuals sum and outlier; IVW: Inverse variance weighted (multiplicative random effects)

## 4. Discussion

To the best of our knowledge, this study is the first comprehensive investigation of the association between PUFAs and cholelithiasis risk based on data from a large-scale observational study combined with MR analysis of large-scale genetic data. In this study, we used the nationally representative NHANES 2017-2020 cohort and two-sample MR to integrate observational studies to investigate the association between dietary intake levels of PUFAs and cholelithiasis. In addition, we used two-step mediated MR to further investigate the association between plasma lipid-mediated Omega-3 levels and the risk of developing cholelithiasis. The results of the cross-sectional observational study showed a negative association between the level of dietary intake of PUFAs and the risk of cholelithiasis, and the two-sample MR approach further confirmed the causal relationship between the level of PUFAs and the development of cholelithiasis. Further, using mediator MR analysis, we found that LDL-C mediated the significance of Omega-3 levels on cholelithiasis occurrence. In particular, the mediating effect of LDL-C was 28%.

Recent MR studies on the causal relationship between Omega-3, Omega-6, and cholelithiasis support our conclusions [21]. However, we did not find a direct causal association between Omga-6 and cholelithiasis in our MR analysis because we analyzed the data using the most recent, larger sample of GWAS data available than in previous studies, avoiding false-positive results that might have resulted from small sample data in previous studies, and using various sensitivity analyses to rule out the possibility of bias from horizontal pleiotropy.

Plasma LDL-C concentrations are considered to be an important factor in the development of cardiovascular disease, but the LDL-C that contributes to this risk is largely related to its particle size. Subsequent large cohort studies have found that only larger particles of LDL concentration and mean particle size were positively associated with Omega-3 intake, in contrast to smaller particles of LDL concentration, which were not [22, 23]. Omega-3 polyunsaturated fatty acid (PUFA) supplementation may have increased LDL particle size by decreasing Apo b100 concentrations without decreasing LDL cholesterol levels [24]. In terms of clearance of VLDL, the increased metabolism of VLDL by Omega-3 may imply a high conversion to larger LDL particles [22]. Importantly, larger particle density LDL-C is considered to be protective against CVD in cardiovascular disease. In previous analyses of populations, it was found that a larger average LDL size was associated with a lower risk of future CVD [25, 26]. From the most recent prospective cohort studies, plasma LDL-C and cholelithiasis risk are negatively correlated, with a decrease in LDL-C elevating cholelithiasis risk. Similarly, a growing body of evidence from MR analyses suggests that genetically reduced plasma LDL-C increases cholelithiasis risk [13, 27]. And, there is evidence that this association may be linear [27].

Our study also has some limitations. Firstly, after correcting for all covariates, the relationship between the level of dietary intake of PUFAs and the development of cholelithiasis became less significant, with only PUFA20:4 remaining nominally statistically significant (p=0.088). The likely reason for this is that cholelithiasis, as a complex disease of the biliary system whose specific etiology is not yet clear, may be associated with other relevant underlying disease conditions and metabolic factors in its development, and adjusting for confounding effects such as diabetes, hypertension, and hyperlipidemia at the baseline level may have only moderately reduced their impact on the observed results. Importantly, MR analyses performed subsequently provided a higher level of evidence for the study, and we therefore concluded that PUFAs, particularly Omega-3, have an important role in the development of cholelithiasis. In addition, because the main population in the observational study cohort and the MR analysis cohort was of European descent and few participants of Asian ethnicity were included, the conclusions may not be representative of all populations and cannot be fully generalized to other ethnic populations, including those of Asian descent. In addition, due to the lack of relevant data, only the Omega-3, and Omega-6 major categories were analyzed as subgroups of PUFAs in our MR study, without encompassing all individually classified PUFAs, which may result in the association between one of these determinants and the development of cholelithiasis not being observed individually, which is expected since the MR study used publicly available GWAS data from external sources, recent developments in GWAS may help to expand this part of the study.

In conclusion, our findings suggest that PUFAs may act as protective factors for the development of cholelithiasis, in which Omega-3 reduces the risk of cholelithiasis by mediating a rise in LDL-C levels and thereby reducing the risk of cholelithiasis. In clinical practice, people without cholelithiasis should be encouraged to consume diets or related supplements containing PUFAs in moderation to reduce the risk of cholelithiasis.

## 5. Author contributions

Y.Z conceived and designed the current study., Y.C and W.C performed analyses. Y.C and J.Q prepared the manuscript. X.J and C.W reviewed and edited the manuscript. All authors read and approved the manuscript.

## 6. Funding information

This work was supported by the National Key Clinical Specialty (General Surgery) of the First Affiliated Hospital of Wenzhou Medical University.

## 7. Conflict of Interest

The authors declare no competing interests.

## 8. Data availability

Publicly available summary statistics are obtained from https://gwas.mrcieu.ac.uk/ (IEU open GWAS), https://www.cdc.gov/nchs/nhanes/index.htm (NHANES), https://www.ukbiobank.ac.uk/ (UK biobank), https://csg.sph.umich.edu/willer/public/glgc-lipids2021/ (GLGC) and https://www.finngen.fi/en (FinnGen).

## 9. Acknowledgements

This research was conducted using data from the Genome-wide association study(GWAS)summary statistics from the public website “IEU open GWAS”, FinnGen, UKbiobank, and Global Lipids Genetics Consortium. We thank the participants, contributors, clinicians, and researchers for making data available for this study.

---

**Supplementary table 1:** Cochrane’s Q test result.

| **Exposure** | **Outcome** | **Method** | **Q** | **Q_df** | **Q_pval** |
| --- | --- | --- | --- | --- | --- |
| PUFA | cholelithiasis | MR Egger | 1769.71 | 51.00 | 0.00E+00 |
| PUFA | cholelithiasis | Inverse variance weighted | 1783.10 | 52.00 | 0.00E+00 |
| MUFA | cholelithiasis | MR Egger | 626.34 | 53.00 | 5.95E-99 |
| MUFA | cholelithiasis | Inverse variance weighted | 665.67 | 54.00 | 2.88E-106 |
| Omega-3 | cholelithiasis | MR Egger | 312.47 | 43.00 | 1.37E-42 |
| Omega-3 | cholelithiasis | Inverse variance weighted | 313.89 | 44.00 | 2.01E-42 |
| Omega-6 | cholelithiasis | MR Egger | 1831.88 | 51.00 | 0.00E+00 |
| Omega-6 | cholelithiasis | Inverse variance weighted | 1834.59 | 52.00 | 0.00E+00 |
| Omega-3 | HDL-C | MR Egger | 18809.98 | 46.00 | 0.00E+00 |
| Omega-3 | HDL-C | Inverse variance weighted | 18810.12 | 47.00 | 0.00E+00 |
| Omega-3 | LDL-C | MR Egger | 13244.94 | 46.00 | 0.00E+00 |
| Omega-3 | LDL-C | Inverse variance weighted | 14388.63 | 47.00 | 0.00E+00 |
| Omega-3 | TG | MR Egger | 28467.52 | 46.00 | 0.00E+00 |
| Omega-3 | TG | Inverse variance weighted | 30577.29 | 47.00 | 0.00E+00 |
| LDL-C | cholelithiasis | MR Egger | 5519.47 | 299.00 | 0.00E+00 |
| LDL-C | cholelithiasis | Inverse variance weighted | 5519.74 | 300.00 | 0.00E+00 |
| TG | cholelithiasis | MR Egger | 2278.89 | 345.00 | 1.69E-281 |
| TG | cholelithiasis | Inverse variance weighted | 2310.85 | 346.00 | 5.47E-287 |

**Supplementary table 2:**
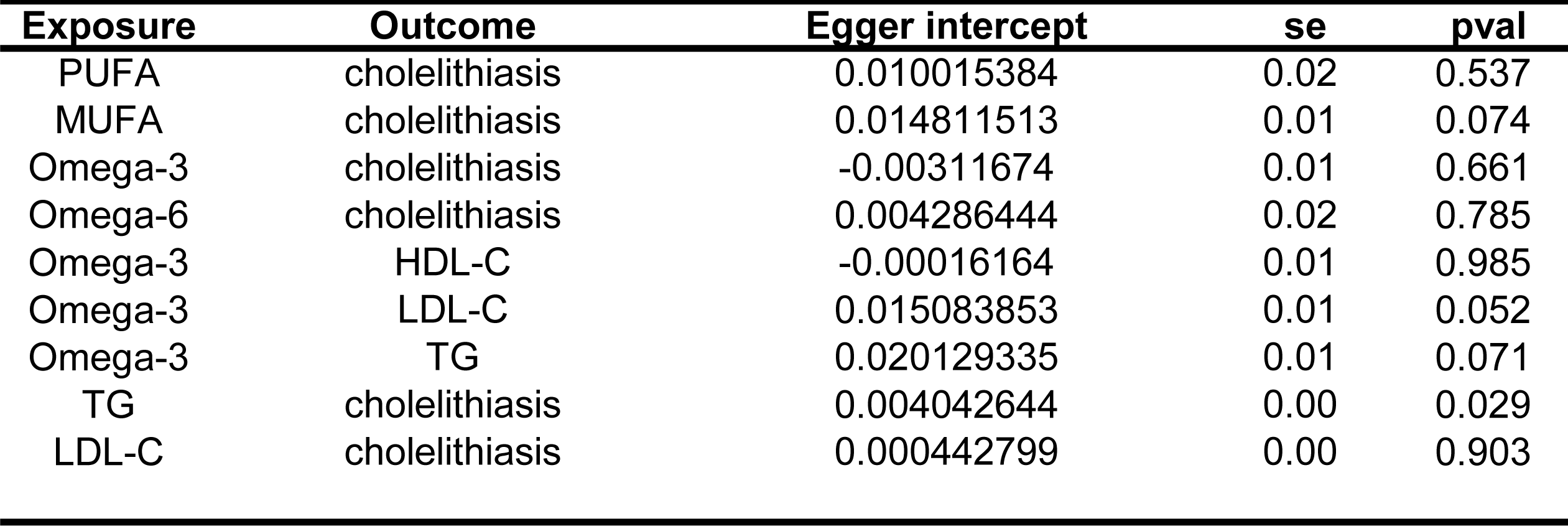
MR Egger’s intercept test.

| **Exposure** | **Outcome** | **Egger intercept** | **se** | **pval** |
| --- | --- | --- | --- | --- |
| PUFA | cholelithiasis | 0.010015384 | 0.02 | 0.537 |
| MUFA | cholelithiasis | 0.014811513 | 0.01 | 0.074 |
| Omega-3 | cholelithiasis | -0.00311674 | 0.01 | 0.661 |
| Omega-6 | cholelithiasis | 0.004286444 | 0.02 | 0.785 |
| Omega-3 | HDL-C | -0.00016164 | 0.01 | 0.985 |
| Omega-3 | LDL-C | 0.015083853 | 0.01 | 0.052 |
| Omega-3 | TG | 0.020129335 | 0.01 | 0.071 |
| TG | cholelithiasis | 0.004042644 | 0.00 | 0.029 |
| LDL-C | cholelithiasis | 0.000442799 | 0.00 | 0.903 |

## Notes

### Competing Interest Statement

The authors have declared no competing interest.

### Summary of Updates

1. Figure 3, which was missed in the last commit, was added. 2. Author information has been updated.

## References

1. Simopoulos, A.P., Essential fatty acids in health and chronic disease. American Journal of Clinical Nutrition, 1999. 70(3): p. 560S–569S.

2. Djuricic, I. and P.C. Calder, Beneficial Outcomes of Omega-6 and Omega-3 Polyunsaturated Fatty Acids on Human Health: An Update for 2021. Nutrients, 2021. 13(7).

3. Cohen, B.I., et al., Dietary fat and fatty acids modulate cholesterol cholelithiasis in the hamster. Lipids, 1992. 27(7): p. 526–32.

4. Kim, J.K., et al., N-3 polyunsaturated fatty acid attenuates cholesterol gallstones by suppressing mucin production with a high cholesterol diet in mice. Journal of Gastroenterology and Hepatology, 2012. 27(11): p. 1745–1751.

5. Tsai, C.J., et al., The effect of long-term intake of cis unsaturated fats on the risk for gallstone disease in men - A prospective cohort study. Annals of Internal Medicine, 2004. 141(7): p. 514–522.

6. Misciagna, G., et al., Diet, physical activity, and gallstones - a population-based, case-control study in southern Italy. American Journal of Clinical Nutrition, 1999. 69(1): p. 120–126.

7. Lee, J.M., et al., Fatty Acid Desaturases, Polyunsaturated Fatty Acid Regulation, and Biotechnological Advances. Nutrients, 2016. 8(1).

8. Berr, F., et al., Dietary N-3 polyunsaturated fatty acids decrease biliary cholesterol saturation in gallstone disease. Hepatology (Baltimore, Md.), 1992. 16(4): p. 960–7.

9. Therien, A., et al., Omega-3 Polyunsaturated Fatty Acid: A Pharmaco-Nutraceutical Approach to Improve the Responsiveness to Ursodeoxycholic Acid. Nutrients, 2021. 13(8).

10. Jang, S.I., et al., Combination treatment with n-3 polyunsaturated fatty acids and ursodeoxycholic acid dissolves cholesterol gallstones in mice. Scientific Reports, 2019. 9.

11. Berr, F., et al., Effect of dietary n-3 versus n-6 polyunsaturated fatty acids on hepatic excretion of cholesterol in the hamster. Journal of lipid research, 1993. 34(8): p. 1275–84.

12. Balasubramaniam, S., et al., Reduction in plasma cholesterol and increase in biliary cholesterol by a diet rich in n-3 fatty acids in the rat. Journal of lipid research, 1985. 26(6): p. 684–9.

13. Chen, L., et al., Insights into modifiable risk factors of cholelithiasis: A Mendelian randomization study. Hepatology, 2022. 75(4): p. 785–796.

14. Smith, G.D. and S. Ebrahim, ’Mendelian randomization’: can genetic epidemiology contribute to understanding environmental determinants of disease? International Journal of Epidemiology, 2003. 32(1): p. 1–22.

15 . Davey Smith, G., N. Timpson, and S. Ebrahim, Strengthening causal inference in cardiovascular epidemiology through Mendelian randomization. Annals of Medicine, 2008. 40(7): p. 524–541.

16. Graham, S.E., et al., The power of genetic diversity in genome-wide association studies of lipids. Nature, 2021. 600(7890): p. 675-+.

17. Lawlor, D.A., et al., Mendelian randomization: Using genes as instruments for making causal inferences in epidemiology. Statistics in Medicine, 2008. 27(8): p. 1133–1163.

18. Bowden, J., G.D. Smith, and S. Burgess, Mendelian randomization with invalid instruments: effect estimation and bias detection through Egger regression. International Journal of Epidemiology, 2015. 44(2): p. 512–525.

19. Bowden, J., et al., Consistent Estimation in Mendelian Randomization with Some Invalid Instruments Using a Weighted Median Estimator. Genetic Epidemiology, 2016. 40(4): p. 304–314.

20. Verbanck, M., et al., Detection of widespread horizontal pleiotropy in causal relationships inferred from Mendelian randomization between complex traits and diseases. Nature Genetics, 2018. 50(5): p. 693-+.

21. Sun, Q., N. Gao, and W. Xia, Association between omega-3/6 fatty acids and cholelithiasis: A mendelian randomization study. Frontiers in Nutrition, 2022. 9.

22. Amigo, N., et al., Habitual Fish Consumption, n-3 Fatty Acids, and Nuclear Magnetic Resonance Lipoprotein Subfractions in Women. Journal of the American Heart Association, 2020. 9(5).

23. Annuzzi, G., et al., Lipoprotein subfractions and dietary intake of n-3 fatty acid: the Genetics of Coronary Artery Disease in Alaska Natives study. American Journal of Clinical Nutrition, 2012. 95(6): p. 1315–1322.

24. Yanai, H., et al., An Improvement of Cardiovascular Risk Factors by Omega-3 Polyunsaturated Fatty Acids. Journal of clinical medicine research, 2018. 10(4): p. 281–289.

25. Mora, S., et al., Lipoprotein Particle Profiles by Nuclear Magnetic Resonance Compared With Standard Lipids and Apolipoproteins in Predicting Incident Cardiovascular Disease in Women. Circulation, 2009. 119(7): p. 931–U44.

26. Mora, S., R.J. Glynn, and P.M. Ridker, High-Density Lipoprotein Cholesterol, Size, Particle Number, and Residual Vascular Risk After Potent Statin Therapy. Circulation, 2013. 128(11): p. 1189–1197.

27. Chen, L., et al., Novel insights into causal effects of serum lipids and lipid-modifying targets on cholelithiasis. Gut, 2024. 73(3): p. 521–532.

